# Family and provider perceptions of quality of care in the management of sick young infants in primary health care settings in four counties of Kenya

**DOI:** 10.1101/2020.07.20.20158428

**Authors:** Samuel Mungai Mbugua, Jesse Gitaka, Tabither Gitau, George Odwe, Peter Mwaura, Wilson Liambila, Charity Ndwiga, Kezia K’Oduol, Charlotte E. Warren, Timothy Abuya

## Abstract

**Background:** Understanding the perceptions of quality of care given to sick young infants in primary health care settings is key for developing strategies for effective uptake and utilization of PSBI guidelines. The purpose of this study is to assess families’ and providers’ perceptions of care given to sick young infants at primary healthcare facilities in four diverse counties in Kenya.

**Methods:** A cross-sectional qualitative design involving in-depth interviews (23) and focus group discussions (25) with very young (15-18 years), young (19-24 years) and older (25-45 years) caregivers of young infants 0-59 days; and key informant interviews with community- and facility-based frontline health providers (14) in primary health care facilities. Qualitative data were captured using audio tapes and field notes, transcribed, translated, and exported into QSR NVivo 12 for analysis. A thematic framework approach was adopted to classify and analyze data.

**Results:** Perceived care given to SYIs was described around six domains of WHO’s framework for the quality of maternal and newborn health care: evidence-based practices for routine and emergency care; functional referral systems; effective communication; respect and preservation of dignity; availability of competent, motivated human resources; and availability of physical resources. Views of caregivers and providers regarding SYIs care at PHCs were similar across the four sites. Main hindrance to SYI care includes stockout of essential drugs, limited infrastructure, lack of functional referral system, inadequate providers which led to delays in receiving treatment, inadequate provider skills and poor provider attitudes. Despite these challenges, motivation and teamwork of health providers were key tenets in care provision.

**Conclusion:** The findings underscore the need to prioritize improving quality of SYIs services at PHCs by building capacity of providers through training, ensuring continuous supply of essential medicines and equipment, improving infrastructure including referral.

## Background

Recent estimates show a substantial reduction in global Neonatal Mortality Rates (NMR) by 51% from 36·6 deaths per 1000 livebirths in 1990, to 18·0 deaths per 1000 livebirths in 2017[1]. Despite the decline, over 2.5 million children died in the first month of life in 2018 which translates to approximately 7000 newborn deaths every day[1]. Most of these deaths are attributed to preventable causes such as newborn infections, often due to lack of access to quality of care during birth and the first days of life [2].

The Sustainable Development Goals (SDGs) aim to reduce neonatal mortality to at least 12 deaths per 1,000 live births and under five mortality to 25 deaths per 1,000 live births by 2030 [3,4]. In 2015, the World Health Organization (WHO) developed guidelines for management of possible serious bacterial infection (PSBI) in neonates (0–28 days old) and young infants (0–59 days old) where referral is not feasible. While increasing hospital-based treatment is imperative, if effective treatment for young infants with serious infection could be provided at Primary Health Care (PHC) facilities when families do not accept or are unable to access referral care, many more infants could receive potentially lifesaving care [5].

Kenya has incorporated the WHO’s PSBI guidelines as part of the Integrated Management on Neonatal and Childhood Illness (IMNCI) [6]. This initiative is expected to improve treatment of sick young infants (SYIs) with serious infection at the PHC facilities when families do not accept or are unable to access referral services[5]. However, evidence shows that health systems in resource-limited settings remain unresponsive to provision of optimal quality newborn care [7–9]. Challenges including lack of appropriately trained staff, incorrect treatment, poor staff attitude, delay in referral, poor cooperation and interpersonal relationships between health providers as well as inadequate supplies and equipment may hinder provision of quality care to SYIs [10–12]. Perceptions about the quality of care as judged by users may influence care seeking and subsequent utilization of health services [13–16].

This paper explores caregivers and provider perceptions of quality of care given to SYI at PHCs (dispensaries and health centers) in four diverse counties in Kenya. We draw on WHO’s framework for the quality of maternal and newborn health care[17]. The concept of quality of maternal and newborn care entails two important, inter-linked elements of provision and experience of care by users. In this study, we focus on both domains of the quality at PHCs as perceived by users and providers of health care services.

## Methods

### Study design

We used a cross-sectional qualitative study design. The study draws on data from a formative assessment that is part of implementation research (IR) aimed at guiding the operationalization of PSBI guidelines in Kenya.

### Study setting

Data was collected in four purposively sampled counties. These sites are representative of a mix of varying contexts characterized by rural and urban-slum disadvantage, nomadic pastoralist and agrarian settings that impacts access to health care. The settings have higher infant and neonatal mortality rates than the national mean with many other deaths in the community going unreported. Two sub counties in each county were selected in consultation with respective County Health Management Teams (CHMT). Six facilities in each sub county were subsequently purposively selected as implementation sites. For purposes of presenting the results, we anonymized the sites using symbols as county A, B, C and D.

### Data collection

Caregivers were selected based on age; residency in the project site and with newborns or young infants aged 0-59 days. They were recruited with help of village elders or community health volunteers (CHVs). The interviews were conducted in Kiswahili or local languages by research assistants with training in qualitative data collection using an interview guide. Health providers were interviewed to examine facility level perceptions of quality of care for SYI and challenges faced during service delivery among other aspects. Table 1 outlines type and number of qualitative interviews conducted.

**Table 1:** Distribution of qualitative interviews by site.

| Category of participants | County D | County C | County B | County A | Total |
| --- | --- | --- | --- | --- | --- |
| IDI Very young mothers (15-18 years) | 2 | 4 | 3 | 2 | 11 |
| IDI young mothers (19-24 years) | 3 | 2 | 3 | 4 | 12 |
| IDI with Providers & facility managers | 2 | 2 | 3 | 7 | 14 |
| FGD Very young mothers (15-18 years) | 2 | 1 | 2 | 2 | 7 |
| FGD young mothers (19-24 years) | 3 | 3 | 2 | 4 | 12 |
| FGD Older mothers (25-45 years) | 1 | 1 | 2 | 2 | 6 |
| FGD married men (>35 years) | 2 | 2 | 1 | 2 | 7 |
| FGD with active CHVs | 2 | 2 | 1 | 2 | 7 |

### Data Analysis

Qualitative data were captured via audio tapes and field notes, translated, transcribed and exported into QSR NVivo 12 for analysis. A thematic framework approach was adopted to classify and organize data into themes. We used an iterative analysis process to develop a coding framework. Analysis charts were developed for each theme and categorized across participants and sites in accordance with WHO’s standard for improving quality of maternal and newborn care in health facilities[17].

### Patient and Public Involvement

Community and public engagement activities included community advisory forums and community education days in which sensitization, understanding and dialogue around research priorities was fostered.

## Results

Perception of quality of care were structured around WHO’s domains of provision and experience of care [18]. Under the provision of care, two elements of quality care are presented and include: evidence-based practices for routine care and management of complications, and functional referral systems. In terms of experience of care, three elements of quality care include effective communication, respect and preservation of dignity. We also explored two other quality elements cutting across both domains (availability of competent, motivated human and physical resources).

### a) Timely evidence-based practices for routine care and management of SYI

Most Caregivers were concerned about delays in receiving treatment for SYI at PHCs. The delays were mainly attributed to inadequate number of providers, which sometimes is aggravated by frequent absenteeism or delays in reporting to workstations. In some PHCs, there was only one provider, usually a nurse, who was responsible for everything. As a result, many caregivers experienced long queues and waiting time before their SYI received treatment.

> *“*…*the doctor is just one at the hospital and the patients are many, there is nowhere else to go so we are forced to just queue here”* **FGD, Very Young mothers, 15-18 years, County A**.

Most providers acknowledged that there were limited number of staff available in the health centers and dispensaries which affected delivery of quality care services for the newborns. The shortage of staff contributed to heavy workload and staff burnout which compromised quality of care for newborns.

> *“We have inadequate personnel. To some extent, I think the care given to patients may not met their expectation. Also, staff burnout is common. If a staff has a burnout, s/he may not exhaustively deliver unlike when the work is well distributed to the staff”* **IDI, Facility provider, County D**.

The delays in receiving treatment at PHC is compounded by fragmentation of service architecture or lack of prioritization for SYI as narrated by one community health worker.

> *“Then again when a mother has been referred, they are not given priority to the facility they go to, and so they have to move around from one department to another buying files and registering and all this time she is moving around with the baby who has not yet been treated”*. **FGD, CHV, County B**.

### b) Functional referral systems

Caregivers observed that most facilities did not have ambulances, lacked fuel or necessary equipment and supplies. Providers concurred that some of the facilities did not have ambulance that meet the standards for emergencies, including referrals. The referral challenges are compounded by long distance and poor roads as explained below.

> *“Our vehicle is not up to the standard for an ambulance, it is a matatu. We need an ambulance where there is oxygen that can resuscitate the patient and we need drivers who are trained to provide emergency services*… *Facilities are quite far from the communities making referrals quite challenging*.*”* **IDI, Health provider, County C**.

### c) Effective communication, respect and dignity

A positive client-provider interaction is a key element of quality of care. Some caregivers were satisfied with the information they received from providers about SYIs conditions, procedures required and advice on care.

> *“They*[*providers*] *attended to me very well. I explained to them the problem the baby had. I was told the baby did not have the urge to breastfeed. I was advised to go and buy a cup so I may express the breast milk and feed the baby. The providers even prescribed to me the medicine*.*”* **IDI, Very Young mothers, 15-18 years, County B**.

Caregivers expressed how they were attended to by healthcare providers. Some caregivers expressed being handled very well and receiving quality SYIs services. Others reported feeling belittled when they sought care at facilities claiming that some providers were extremely harsh; would quarrel, abuse or look down upon them. Disrespect on the part of providers discouraged care seeking for SYIs.

> *“*… *you can be explaining to the doctor how the baby is feeling and then s/he starts quarrelling; ‘oh why didn’t you bring the baby earlier than this’ and so many things. They harass us and if they become like this, we may start fearing them and hide when the baby is sick. So, they should stop all these”* **FGD, Very Young mothers, 15-18 years, County B**.

### d) Availability of competent, motivated human resources

For effective and efficient delivery of services, healthcare providers need support; training and motivation to enable them to perform effectively. We present two interrelated themes that illuminate what may hinder or enhance performance of providers as they manage SYI in PHC facilities-providers’ knowledge and passion for work.

#### i) Provider knowledge

When asked about the providers’ knowledge and skills, caregivers implied that the providers lacked effective specialized knowledge and or basic skills necessary to support management of SYI;

> “*Indeed, I have observed that our providers here do not have special skills for infants, they do routine work that applies to both adults and children”* **FGD, Married men >35 years, County D**.

Majority of the respondents across all counties stated that when SYIs are taken for treatment, the providers just prescribe medication without carrying out the necessary laboratory tests. To validate this view, providers were asked the greatest challenge they face while managing SYI and some reported the delicate nature of infants made it hard to have definitive diagnosis as one provider noted: *“Managing an infant is quite challenging because for one thing they are delicate, so in most cases for example if they have a bacterial infection, or if you suspect bacterial infections may be they present with fever, but we usually rule out malaria but now if the baby persist with fever may be you have checked out the cord, you have ruled out tetanus, it’s difficult to come out with the right diagnosis. Basically, managing infants is quite challenging”* **IDI, Health Provider, County B**.

#### ii) Passion for work and teamwork

Most of the providers reported that despite staff shortage, there are occasions where teamwork becomes a key pillar of care giving where providers from various departments support other departments making it easier to manage cases as one provider noted:

> “*Mmm*….*what motivates me is one, the passion for work*…*it is not the salary*…*if it is about the salary I would be coming at eight and leave at four, because am just looking for the salary, so it is the passion for the work that I just want to see that mother, that sick child, tomorrow we meet in the streets I see them healthy, that is what motivates me. The second thing that motivates me is the teamwork spirit that we have in the facility*. **IDI, Health Provider, County C**.

### e) Availability of essential physical resources

Most caregivers reported that some PHC facilities did not have adequate or functional equipment. Caregivers had limited options but to buy drugs from private pharmacies or travel long distances to get testing services.

> *“*… *you find that there are no drugs in the facility, so you are given the prescription to go and buy the drugs from a chemist outlet*.” **FGD, Young mothers 19-24 years, County D**.

The unavailability of drugs in facilities leads to clients resorting to alternative treatments including traditional medicines.

> *“Most of us go for traditional medicine because there are no drugs at the facility so when we come here and find no drugs we still go back to traditional medicine because we have no option*….*”* **FGD, Older men, County A**.

Caregivers also decried of lack of adequate space for managing SYIs in some facilities.

> *“They need to expand the hospital to increase space, sometimes it’s usually very congested, you can even lack space and we cannot access other hospital because they are far*. “**FGD, Young Mothers, 19-24 years, County C**.

Providers pointed to an inability to handle some conditions at the primary health facility level, due to lack of equipment, essential drugs and supplies.

> *“Many times, cases that we could have handled here, we have to refer to a higher level because of lack of essential commodities*. **IDI, Health Provider, County D**.

## Discussion

Quality of care has been recognized as a critical aspect of improving maternal and newborn health outcomes. A high coverage combined with improved quality of care contribute to reduction of maternal and newborn morbidity and mortality[19, 20]. The study contributes to a complex issue of quality of neonatal care in low-resource settings by examining perspective of caregivers and providers. Perceptions on quality of care, rather than clinical indicators of quality drives utilization of health services and are essential to increasing demand[21].

Our finding illustrate commonality between caregivers and providers on the need to improve competency of providers to manage SYIs. Gaps in provider skills and competencies have huge implications for managing SYI and compromise technical quality of care. This means that providers require frequent updates on management of SYI, an important observation documented in IMNCI implementation which shows that continuous training was essential for improving quality of care in child health services [22]. Enhancing provider knowledge through initiatives such as mentorship, on-job training and online approaches to build their confidence in providing SYI care is essential. A systematic review of effectiveness of capacity building interventions relevant to public health practice reported six intervention types: internet-based instruction, training and workshops, technical assistance, education using self-directed learning, communities of practice, and multi-strategy interventions. The review shows that organizations should carefully consider methods of capacity building interventions based on purpose. [23].

Providers on the other hand were concerned about the number of staff available and equipment available for them to work more efficiently and deliver optimum care to SYI at PHC. Low staff numbers lead to increased workload reported previously [24]. Our study revealed that inadequacy of drugs and equipment, infrastructure and space as major barriers towards effective service delivery for SYI at PHCs. These features lead to delays in treatment, especially where referral systems are weak or lacking [25]. Most facilities did not have working ambulances, further creating additional burden to caregivers whose babies required referral service. For effective adoption of PSBI, strategies to equip lower level PHC with essential equipment and drugs will ensure timely access to treatment for SYI and reduce unnecessary referrals to higher level facilities a phenomenon that has been documented elsewhere [26, 27].

Respectful and dignified care has also been recognized as a key component of quality of newborn care during postnatal period [28–31], however documentation of experiences of mistreatment among infants is limited [32]. Our study found that some caregivers complained of disrespectful treatment by providers, negative provider attitudes, and lack of empathy, which are deterrents to care-seeking. Although we did not examine deeply the extent in which this deters care seeking for SYI, elsewhere strategies to improve interaction with caregivers have demonstrated success [33].

Other interlinked elements of care that requires attention is delayed initiation of treatment due to a range of factors including provider competency in diagnosing PSBI, long queues emanating from inadequate number of health providers, poor triaging process, provider attitude as well as nepotism and discrimination of caregivers due to their socio-economic status. The complex interaction of these issues means that simple strategies such as sensitizing providers for positive provider-client interaction has the potential to improve PSBI uptake by caregivers. Overall, provision of efficient, equitable, compassionate, reliable, timely, patient-centered care while applying evidence-based standards to ensure the safety of clients and provider’s satisfaction [29–31], are key elements of care that need to be incorporated in scaling up PSBI strategies.

## Conclusion

Understanding caregivers’ and providers’ perceptions of quality of care given to SYI, is critical in the roadmap towards ensuring effective uptake and utilization of PSBI guidelines in the management of SYI within the existing IMNCI strategies. The findings clearly point to the areas that need prioritization to improve the quality of SYIs health services. They include a need for continuous skill and knowledge enhancement of providers who manage SYIs, continuous supply of essential medicines and equipment in the facility and the need for service arrangement strategies in the context of constrained infrastructure. This will reduce the gaps of optimal quality for care of SYI while ensuring respectful client management, better communication and supportive care. Results from these assessments provide evidence that will be used to support uptake of PSBI in PHC settings. Using an IR approach, providers and managers will use this evidence to localize solutions to improve care for SYI.

## Data Availability

The datasets used and/or analyzed during the current study are available from the corresponding author on reasonable request.

## List of Abbreviations

PSBI: Possible Severe Bacterial Infection
SYI: Sick Young Infants
IMNCI: Integrated Management of Newborn and Childhood Illness
IR: Implementation Research
FGD: Focus Group Discussion
IDI: In Depth Interview
CHV: Community Health Volunteer
PHC: Primary Health Care

## Ethics Approval and consent to participate

This study was approved by the AMREF Ethics and Scientific Review Committee (as ESRC P430/2018), Population Council’s Institutional Review Board (as Protocol 838) and Mount Kenya University Ethics Research Committee. Written informed consent was obtained from each participant before conducting an interview. Assent was attained from participants below the age of majority of 18 years. A written informed consent was also obtained from a parent or guardian of participants below the age of majority. Identifying details were not included during data collection, data entry or analysis.

## Consent for publication

All authors gave their approval and consent for publication of this manuscript.

## Author’s contributions

SM conceptualized the idea, conducted the analysis, and wrote the first draft. GO, TA and CW participated in the interpretation of results and reviewing the manuscript for substantial intellectual content. SM, GO, JG, WL, TA, TG, PM, CN, KO reviewed the manuscript. All authors read and approved the manuscript for publication.

## Acknowledgement

The authors would like to acknowledge all the staff from the participating health facilities and members of the community in the four counties.

The project that generated data used in this study was made possible by the generous support of the American people through the United States Agency for International Development (USAID) under the terms of AID-OAA-A-17-00031. The contents of this manuscript are the sole responsibility of the authors and do not necessarily reflect the views of USAID or the United States Government.

